# Australia’s worsening mental health – what’s next?

**DOI:** 10.1101/2021.07.21.21259430

**Authors:** Joanne Enticott, Shrinkhala Dawadi, Frances Shawyer, Brett Inder, Ellie Fossey, Helena Teede, Seb Rosenberg, Ingrid Ozol, Graham Meadows

## Abstract

**Objectives:** To examine trends in psychological distress in Australia between 2001 to 2017-18, including analysis by age, sex, and location.

**Design, setting and participants:** Secondary analysis of six successive national health surveys of representative samples of the working age population (18-64 years).

**Main outcome measures:** Prevalence of psychological distress at very-high symptom level (defined by a Kessler Psychological Distress Scale [K10] score of 30 or more) and combined high/very-high level (K10 score of 22 or more).

**Results:** The latest survey showed 5.1% of Australians reporting very-high level distress and 14.8% combined high/very-high level - both the largest rates recorded this century. The greatest increase from 2001 to 2017-18 was in women aged 55-64 with very-high distress significantly increasing from 3.5% (95% CI: 2.5-4.5%) to 7.2% (5.9-8.5%), and; high/very-high distress from 12.4% (10.5-14.2%) to 18.7% (16.7-20.7%). Men aged 25-34 had very-high distress increase from 2.1% (1.4-2.8) to 4.0% (2.9-5.1%); and combined high/very-high distress remained stable at 10.6% (9.1-12.1%) to 11.5% (9.7-13.3%). In 2017-18, greatest distress was in women aged 18-24 years (very-high 8.0% (5.9-10.2%); high/very-high 22.1% (18.8-25.3%)). Overall, distress was significantly more prevalent in inner regional Australia than elsewhere (very-high level 4.8% (4.4-5.1%); high/very-high 14.4% (13.8-15%)).

**Conclusions:** Australia’s annual mental health expenditure over this period has doubled, yet population level psychological distress has increased. A whole of government approach and targeted strategies focusing on groups with the poorest mental health such as older working aged women, younger people, particularly women, and those outside of major cities are indicated.

**Box:** **“The known”** Previous examinations of national health surveys had suggested that population mental health was stable as measured by psychological distress.

**“The new”** Examining six consecutive national surveys we provide evidence that mental health has significantly deteriorated between 2001 and 2018. The latest survey showed 5.1% of Australians reporting very-high distress and 14.8% combined high/very-high distress, which are the largest rates reported this century.

**“The implications”** Whole of government approach and targeted strategies focusing on groups with the poorest mental health such as older working aged women, younger people particularly women, and those outside of major cities are indicated.

## 1. Introduction

Health and mental health are influenced by many factors: social determinants link with gender, biological and environmental factors, health and other policies to influence mental health outcomes across the lifespan (1-3). When ill health occurs access to care varies in regional, rural and remote areas and across different communities (4-6). Even when it is accessible, variable educational attainment, income, social status and support, cultural and other factors influence personalised lifestyle and health behaviours, including care-seeking and engagement (3, 7). So mental illness prevalence, access and utilisation of mental health services can vary widely within subgroups.

The Australian National Health Survey (NHS), a regular and invaluable source of data on health and social determinants (8), includes the Kessler-10 (K-10) questionnaire (9). The K10 is a measure of psychological distress that includes anxiety and affective symptoms. High (22-29), and very-high (30-50) K10 band scores are strongly associated with anxiety and affective disorders (9). So regular NHS collection of K10 data enables surveillance of mental disorder trends in the Australian population.

Federal and state governments in Australia have more than doubled constant-dollar per-capita mental health services expenditure since 1992 (4). Around 11% of Australian adults use some mental health service annually (10). If this care was effective, equitably reaching those at greater risk and need, and assuming other influences were stable, improvement would be anticipated. But up to 2014, noting that whole-of-population NHS-K10 trend comparisons may obscure subgroup changes, population NHS K10 scores remained relatively unchanged (4, 11, 12), increasing recently in youth (13). Many other social determinants of health (1) have not in fact been stable, including increasing inequities of income and wealth (14). Concern has been voiced regarding Australia’s mental health service delivery system quality and effectiveness, (eg (12, 13)), service delivery inequity, and possibly forms of iatrogenesis (5).

Building from previous work (11-13), here we examine prevalence of psychological distress in Australia between 2001 and 2018, exploring subgroups by age, gender and location and consider the policy implications arising.

## 2. Methods

### 2.1. Design

This study was a large-scale secondary analysis (n=78, 204) of K10 data from six national data sources collected from working aged Australian adults across the National Health Surveys (NHS) (2001-02, 2004-05, 2007-08, 2011-12, 2014-15, 2017-18), collected by the Australian Bureau of Statistics. We analysed responses from adults aged 18-64 years in each survey, except for the 2004-05 NHS as data was only available for adults aged 20-64 years. We standardised all surveys to the 2001 Australian census population based on the strata of sex and age (15). Elevated psychological distress were calculated, and compared across sex as available in the NHS.

#### National health surveys

The NHS are cross-sectional household-based surveys undertaken at 3-year intervals to monitor health trends over time with detailed methods described elsewhere (8). Trained ABS interviewers conducted face-to-face interviews in each survey. Household and person weights are assigned by the ABS to adjust for the probability of sample selection, seasonality and non-response, and the data are then calibrated to the population benchmarks. This ensures that the estimates are representative of population distributions and compensates for any over-or under-representation of particular categories of persons or households.

#### Psychological distress measure

The K10, a self-administered 10-item Likert scale tool, measures current psychological distress, particularly symptoms of anxiety and depressive disorders (9). Used in ordinal form, band scores are closely associated with mental health disorders (9). K10 scores range between 10 and 50, and score bands are: low (10–15), moderate (16–21), high (22–29) and very-high (30–50). Here we also generated a combined high/very-high category, which consisted of scores 22 and higher.

#### Geographic location

A residential location variable for each survey participant is available and based on the Accessibility and Remoteness Index of Australia (ARIA+) (8). It describes the residential location as Major cities of Australia, Inner Regional Australia or Other.

#### Data analysis

All statistical analyses were performed in Stata 16.0 (StataCorp). When not stratified by age, data were directly age-standardised against the estimated resident population of Australia at 30 June 2001 (15). Effect size estimates for dichotomous outcomes of combined high/very-high and very-high psychological distress are presented as odds ratios calculated using logistic regression on the K10 data from the Australian working age population. Independent variables examined first in a univariate regression with the outcome, then in a multivariable regression, were: year, sex, age-group and location. All independent variables were specified as categorical, including the ‘year’ variable because prevalence changes over time was not linear. For time trend examinations the reference year was 2001. Level of significance for the regressions was set at an alpha of 0.05.

Comparing 2001 and 2017-18 data: Pairwise comparisons using tests for two proportions and a Bonferroni correction for multiple comparisons was applied. Given that twelve sub-group comparisons were planned (see Table 2), to minimise the occurrences of spurious positives, the alpha value was set at 0.0042 (i.e., approx. 0.05/12) for these set of pairwise comparisons.

#### Ethics approval

As is common practice for the ABS, data collection occurred under the auspices of the Census and Statistics Act 1905, and the analyses were approved by both the Australian Parliament and the Privacy Commissioner.

## 3. Results

In the six national surveys between 2001 and 2017-18 there were n=78,204 surveys completed by working age adults producing K10 distress data, see Table 1. Figure 1 shows that the greatest distress occurred in the latest survey at 2017-18: for combined high/very-high level distress the 14.8% rate was significantly greater than all previous years (p<0.001), and; for very-high level distress the 5.1% rate was significantly greater than 2001, 2004, 2007 & 2011 (p<0.01).

**Figure 1:**
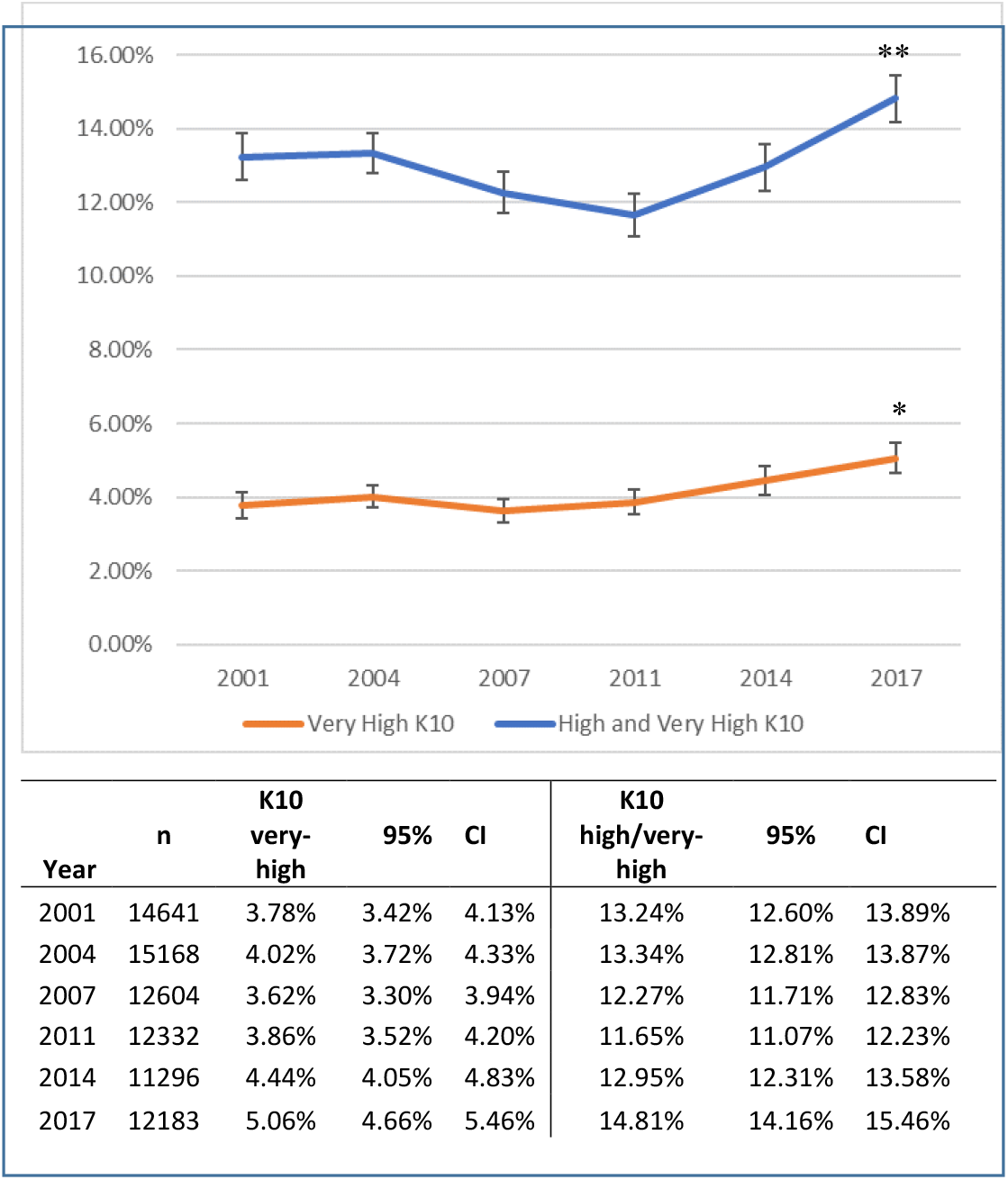
Age-standardised prevalence of psychological distress in the Australian working age population, 2001-2017. ^a^ Standardised to 2001 Australian Census. Derived from a total of n=78,204 survey participants aged 18-64 years. ** Rate at 2017 significantly greater than all previous years (p<0.001). * Rate in 2017 significantly greater than 2001, 2004, 2007 & 2011 (p<0.01).

**Figure 2.**
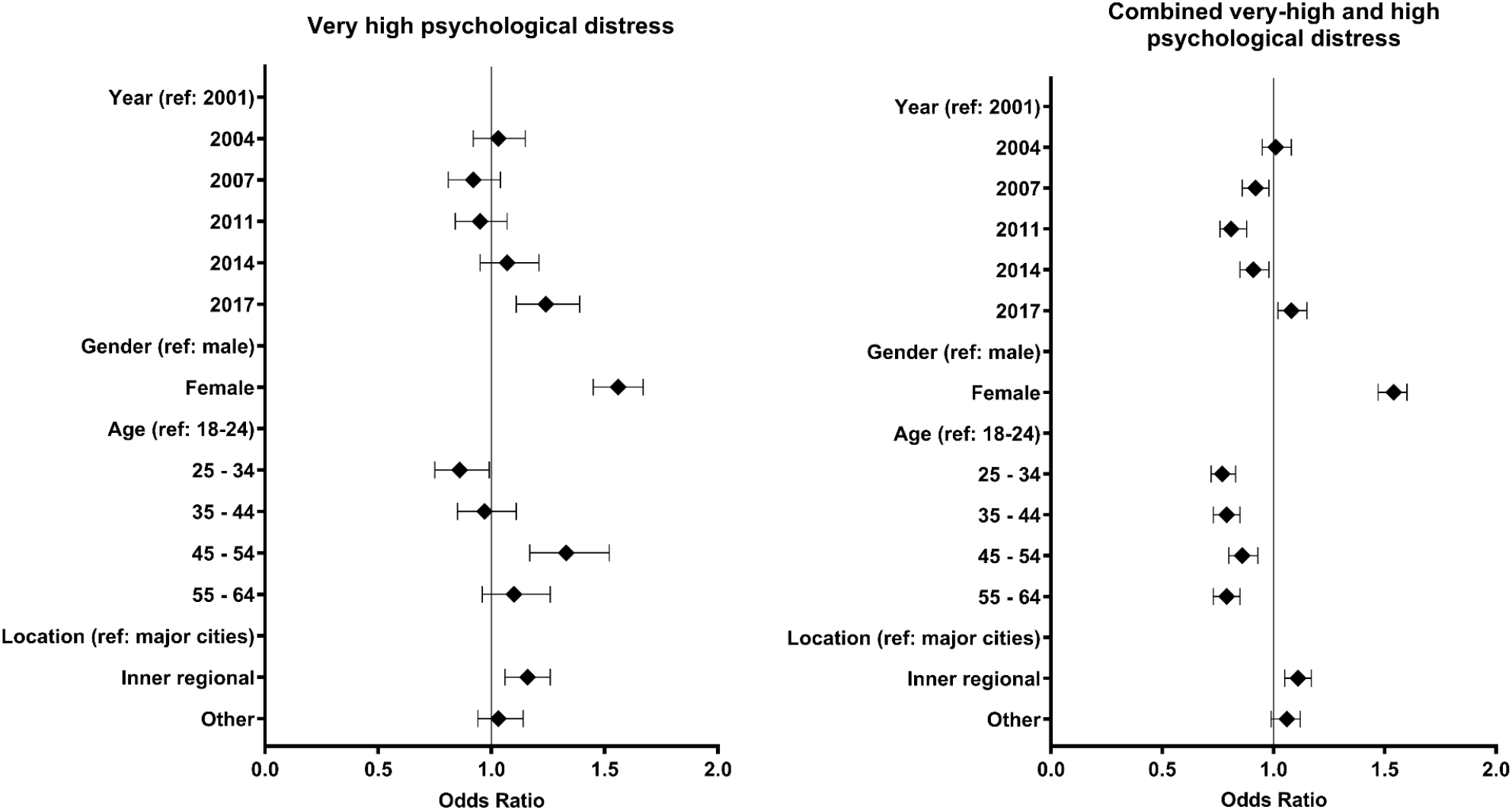
Effect size estimates of psychological distress in the Australian working age population, 2001-2017. Derived using logistic regression on K10 data from n=78,204 adults.

For very-high distress, the lowest rate appeared to be 3.6% (95% Confidence Interval: 3.3-3.9) in 2007; however, confidence intervals (CI’s) overlapped with all other years except 2017-18, where rates appeared significantly greater at 5.1% (95% CI: 4.7-5.5). Multivariable regression confirmed similar rates of very-high distress across 2001, 2004, 2007, 2011 and 2014; and a greater rate in 2017-18 [odds ratios (OR) 1.24, 95% CI: 1.11 to 1.39] compared to 2001, see Table 4.

**Table 3:** Age-standardised prevalence of psychological distress in the Australian working age population, 2001-2017. ^a^ Standardised to 2001 Australian Census. Derived from a total of n=78,204 survey participants aged 18-64 years.

|  | K10 very-high |  |  |  | K10 combined high/very-high |  |  |  |
| --- | --- | --- | --- | --- | --- | --- | --- | --- |
|  | n | Standardised Rate<sup>a</sup> | 95% | CI | n | Standardised Rate<sup>a</sup> | 95% | CI |
| *Age Group* |  |  |  |  |  |  |  |  |
| 18 - 24 | 7846 | 4.05% | 3.62% | 4.49% | 7846 | 15.75% | 14.95% | 16.56% |
| 25 - 34 | 17292 | 3.55% | 3.27% | 3.82% | 17292 | 12.78% | 12.28% | 13.28% |
| 35 - 44 | 19874 | 3.97% | 3.69% | 4.24% | 19874 | 12.99% | 12.52% | 13.46% |
| 45 - 54 | 17742 | 5.39% | 5.05% | 5.72% | 17742 | 14.00% | 13.49% | 14.51% |
| 55 - 64 | 15470 | 4.53% | 4.20% | 4.85% | 15470 | 12.96% | 12.43% | 13.49% |
| *Location<sup>a</sup>* |  |  |  |  |  |  |  |  |
| Major cities | 51289 | 4.09% | 3.92% | 4.27% | 51289 | 13.13% | 12.83% | 13.43% |
| Inner regional | 15073 | 4.75% | 4.40% | 5.10% | 15073 | 14.42% | 13.83% | 15.00% |
| Other | 11862 | 4.24% | 3.87% | 4.61% | 11862 | 13.66% | 13.01% | 14.30% |
| *Sex<sup>a</sup>* |  |  |  |  |  |  |  |  |
| Male | 36809 | 3.30% | 3.12% | 3.49% | 36809 | 10.90% | 10.57% | 11.22% |
| Female | 41415 | 5.18% | 4.96% | 5.41% | 41415 | 16.05% | 15.68% | 16.42% |
| *Male and Year<sup>a</sup>* |  |  |  |  |  |  |  |  |
| 2001 | 6797 | 3.01% | 2.60% | 3.41% | 6797 | 10.86% | 10.11% | 11.61% |
| 2004 | 7135 | 3.27% | 2.86% | 3.69% | 7135 | 11.33% | 10.57% | 12.08% |
| 2007 | 6095 | 2.87% | 2.46% | 3.28% | 6095 | 10.29% | 9.52% | 11.06% |
| 2011 | 5871 | 3.08% | 2.63% | 3.52% | 5871 | 9.54% | 8.77% | 10.31% |
| 2014 | 5197 | 3.43% | 2.92% | 3.93% | 5197 | 10.35% | 9.50% | 11.19% |
| 2017 | 5714 | 4.19% | 3.66% | 4.72% | 5714 | 12.57% | 11.68% | 13.46% |
| *Female and Year<sup>a</sup>* |  |  |  |  |  |  |  |  |
| 2001 | 7844 | 5.19% | 4.69% | 5.69% | 7844 | 17.00% | 16.15% | 17.84% |
| 2004 | 8033 | 5.02% | 4.54% | 5.50% | 8033 | 16.73% | 15.89% | 17.58% |
| 2007 | 6509 | 4.59% | 4.08% | 5.11% | 6509 | 15.33% | 14.44% | 16.23% |
| 2011 | 6461 | 4.61% | 4.09% | 5.13% | 6461 | 13.72% | 12.85% | 14.58% |
| 2014 | 6099 | 5.43% | 4.85% | 6.02% | 6099 | 15.50% | 14.55% | 16.44% |
| 2017 | 6469 | 5.90% | 5.31% | 6.50% | 6469 | 16.97% | 16.03% | 17.92% |

**Table 4:**
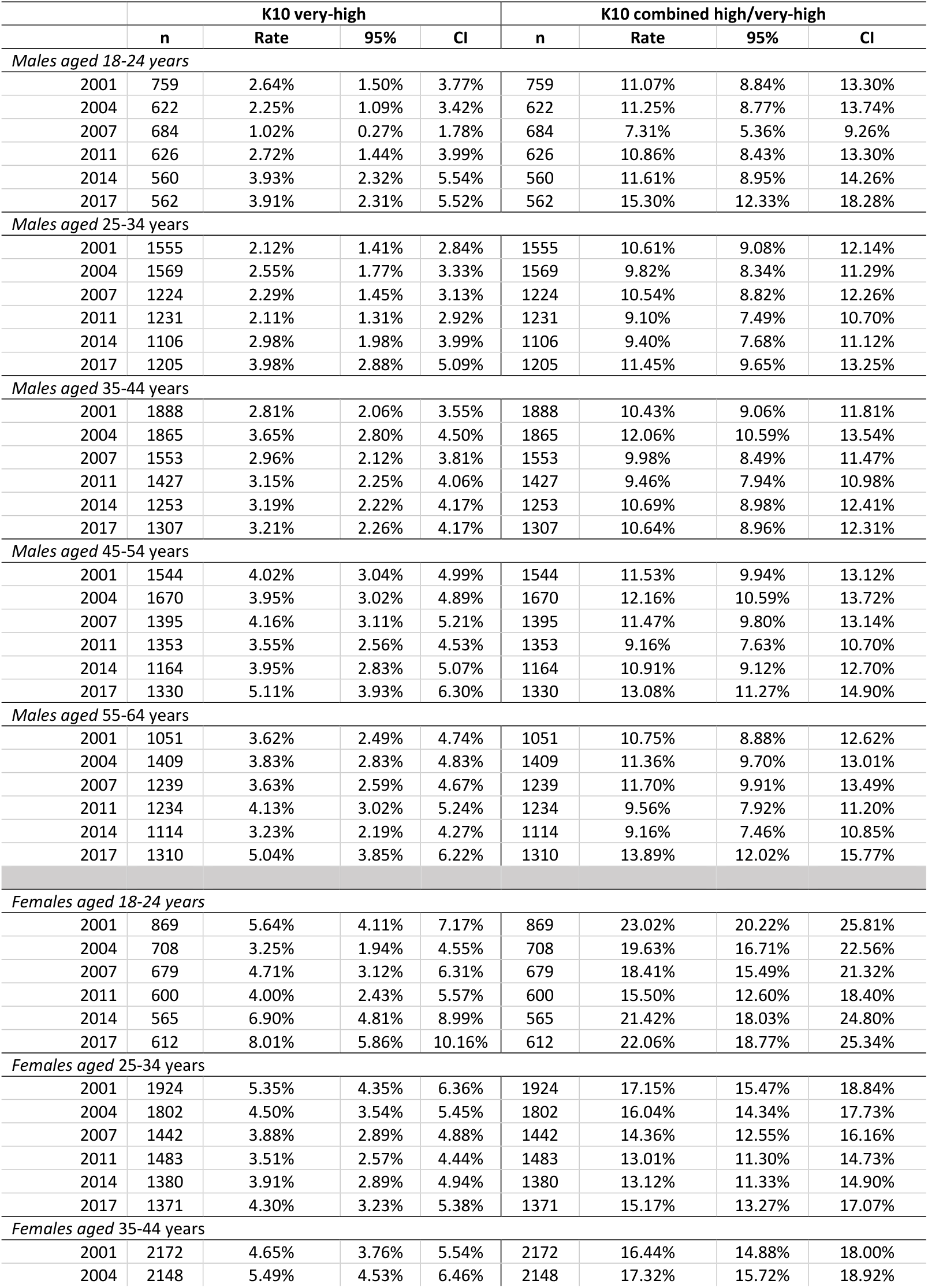
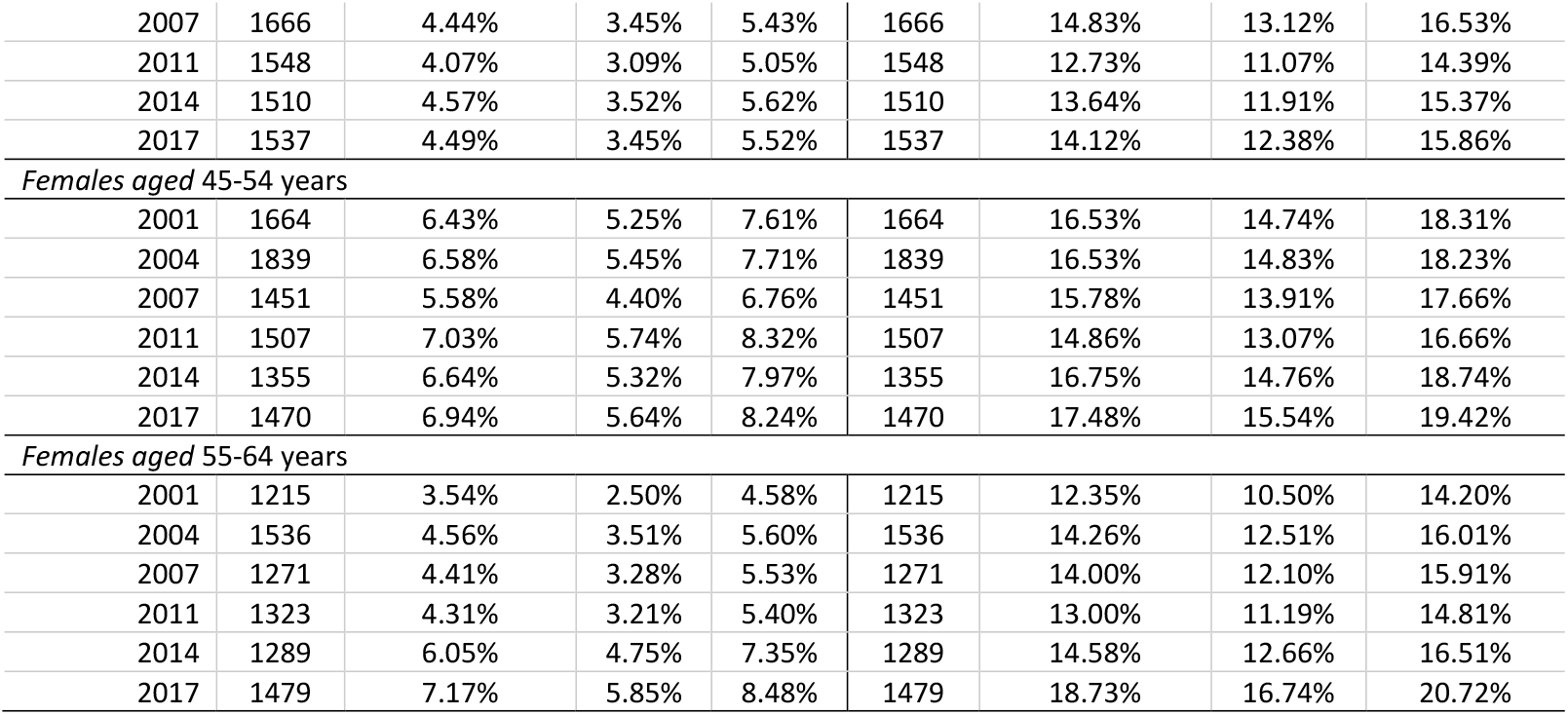
Prevalence of psychological distress by sex and age-groups over time.

|  | K10 very-high |  |  |  | K10 combined high/very-high |  |  |  |
| --- | --- | --- | --- | --- | --- | --- | --- | --- |
|  | n | Rate | 95% | CI | n | Rate | 95% | CI |
| *Males aged 18-24 years* |  |  |  |  |  |  |  |  |
| 2001 | 759 | 2.64% | 1.50% | 3.77% | 759 | 11.07% | 8.84% | 13.30% |
| 2004 | 622 | 2.25% | 1.09% | 3.42% | 622 | 11.25% | 8.77% | 13.74% |
| 2007 | 684 | 1.02% | 0.27% | 1.78% | 684 | 7.31% | 5.36% | 9.26% |
| 2011 | 626 | 2.72% | 1.44% | 3.99% | 626 | 10.86% | 8.43% | 13.30% |
| 2014 | 560 | 3.93% | 2.32% | 5.54% | 560 | 11.61% | 8.95% | 14.26% |
| 2017 | 562 | 3.91% | 2.31% | 5.52% | 562 | 15.30% | 12.33% | 18.28% |
| *Males aged 25-34 years* |  |  |  |  |  |  |  |  |
| 2001 | 1555 | 2.12% | 1.41% | 2.84% | 1555 | 10.61% | 9.08% | 12.14% |
| 2004 | 1569 | 2.55% | 1.77% | 3.33% | 1569 | 9.82% | 8.34% | 11.29% |
| 2007 | 1224 | 2.29% | 1.45% | 3.13% | 1224 | 10.54% | 8.82% | 12.26% |
| 2011 | 1231 | 2.11% | 1.31% | 2.92% | 1231 | 9.10% | 7.49% | 10.70% |
| 2014 | 1106 | 2.98% | 1.98% | 3.99% | 1106 | 9.40% | 7.68% | 11.12% |
| 2017 | 1205 | 3.98% | 2.88% | 5.09% | 1205 | 11.45% | 9.65% | 13.25% |
| *Males aged 35-44 years* |  |  |  |  |  |  |  |  |
| 2001 | 1888 | 2.81% | 2.06% | 3.55% | 1888 | 10.43% | 9.06% | 11.81% |
| 2004 | 1865 | 3.65% | 2.80% | 4.50% | 1865 | 12.06% | 10.59% | 13.54% |
| 2007 | 1553 | 2.96% | 2.12% | 3.81% | 1553 | 9.98% | 8.49% | 11.47% |
| 2011 | 1427 | 3.15% | 2.25% | 4.06% | 1427 | 9.46% | 7.94% | 10.98% |
| 2014 | 1253 | 3.19% | 2.22% | 4.17% | 1253 | 10.69% | 8.98% | 12.41% |
| 2017 | 1307 | 3.21% | 2.26% | 4.17% | 1307 | 10.64% | 8.96% | 12.31% |
| *Males aged 45-54 years* |  |  |  |  |  |  |  |  |
| 2001 | 1544 | 4.02% | 3.04% | 4.99% | 1544 | 11.53% | 9.94% | 13.12% |
| 2004 | 1670 | 3.95% | 3.02% | 4.89% | 1670 | 12.16% | 10.59% | 13.72% |
| 2007 | 1395 | 4.16% | 3.11% | 5.21% | 1395 | 11.47% | 9.80% | 13.14% |
| 2011 | 1353 | 3.55% | 2.56% | 4.53% | 1353 | 9.16% | 7.63% | 10.70% |
| 2014 | 1164 | 3.95% | 2.83% | 5.07% | 1164 | 10.91% | 9.12% | 12.70% |
| 2017 | 1330 | 5.11% | 3.93% | 6.30% | 1330 | 13.08% | 11.27% | 14.90% |
| *Males aged 55-64 years* |  |  |  |  |  |  |  |  |
| 2001 | 1051 | 3.62% | 2.49% | 4.74% | 1051 | 10.75% | 8.88% | 12.62% |
| 2004 | 1409 | 3.83% | 2.83% | 4.83% | 1409 | 11.36% | 9.70% | 13.01% |
| 2007 | 1239 | 3.63% | 2.59% | 4.67% | 1239 | 11.70% | 9.91% | 13.49% |
| 2011 | 1234 | 4.13% | 3.02% | 5.24% | 1234 | 9.56% | 7.92% | 11.20% |
| 2014 | 1114 | 3.23% | 2.19% | 4.27% | 1114 | 9.16% | 7.46% | 10.85% |
| 2017 | 1310 | 5.04% | 3.85% | 6.22% | 1310 | 13.89% | 12.02% | 15.77% |
| *Females aged 18-24 years* |  |  |  |  |  |  |  |  |
| 2001 | 869 | 5.64% | 4.11% | 7.17% | 869 | 23.02% | 20.22% | 25.81% |
| 2004 | 708 | 3.25% | 1.94% | 4.55% | 708 | 19.63% | 16.71% | 22.56% |
| 2007 | 679 | 4.71% | 3.12% | 6.31% | 679 | 18.41% | 15.49% | 21.32% |
| 2011 | 600 | 4.00% | 2.43% | 5.57% | 600 | 15.50% | 12.60% | 18.40% |
| 2014 | 565 | 6.90% | 4.81% | 8.99% | 565 | 21.42% | 18.03% | 24.80% |
| 2017 | 612 | 8.01% | 5.86% | 10.16% | 612 | 22.06% | 18.77% | 25.34% |
| *Females aged 25-34 years* |  |  |  |  |  |  |  |  |
| 2001 | 1924 | 5.35% | 4.35% | 6.36% | 1924 | 17.15% | 15.47% | 18.84% |
| 2004 | 1802 | 4.50% | 3.54% | 5.45% | 1802 | 16.04% | 14.34% | 17.73% |
| 2007 | 1442 | 3.88% | 2.89% | 4.88% | 1442 | 14.36% | 12.55% | 16.16% |
| 2011 | 1483 | 3.51% | 2.57% | 4.44% | 1483 | 13.01% | 11.30% | 14.73% |
| 2014 | 1380 | 3.91% | 2.89% | 4.94% | 1380 | 13.12% | 11.33% | 14.90% |
| 2017 | 1371 | 4.30% | 3.23% | 5.38% | 1371 | 15.17% | 13.27% | 17.07% |
| *Females aged 35-44 years* |  |  |  |  |  |  |  |  |
| 2001 | 2172 | 4.65% | 3.76% | 5.54% | 2172 | 16.44% | 14.88% | 18.00% |
| 2004 | 2148 | 5.49% | 4.53% | 6.46% | 2148 | 17.32% | 15.72% | 18.92% |
| 2007 | 1666 | 4.44% | 3.45% | 5.43% | 1666 | 14.83% | 13.12% | 16.53% |
| 2011 | 1548 | 4.07% | 3.09% | 5.05% | 1548 | 12.73% | 11.07% | 14.39% |
| 2014 | 1510 | 4.57% | 3.52% | 5.62% | 1510 | 13.64% | 11.91% | 15.37% |
| 2017 | 1537 | 4.49% | 3.45% | 5.52% | 1537 | 14.12% | 12.38% | 15.86% |
| *Females aged 45-54 years* |  |  |  |  |  |  |  |  |
| 2001 | 1664 | 6.43% | 5.25% | 7.61% | 1664 | 16.53% | 14.74% | 18.31% |
| 2004 | 1839 | 6.58% | 5.45% | 7.71% | 1839 | 16.53% | 14.83% | 18.23% |
| 2007 | 1451 | 5.58% | 4.40% | 6.76% | 1451 | 15.78% | 13.91% | 17.66% |
| 2011 | 1507 | 7.03% | 5.74% | 8.32% | 1507 | 14.86% | 13.07% | 16.66% |
| 2014 | 1355 | 6.64% | 5.32% | 7.97% | 1355 | 16.75% | 14.76% | 18.74% |
| 2017 | 1470 | 6.94% | 5.64% | 8.24% | 1470 | 17.48% | 15.54% | 19.42% |
| *Females aged 55-64 years* |  |  |  |  |  |  |  |  |
| 2001 | 1215 | 3.54% | 2.50% | 4.58% | 1215 | 12.35% | 10.50% | 14.20% |
| 2004 | 1536 | 4.56% | 3.51% | 5.60% | 1536 | 14.26% | 12.51% | 16.01% |
| 2007 | 1271 | 4.41% | 3.28% | 5.53% | 1271 | 14.00% | 12.10% | 15.91% |
| 2011 | 1323 | 4.31% | 3.21% | 5.40% | 1323 | 13.00% | 11.19% | 14.81% |
| 2014 | 1289 | 6.05% | 4.75% | 7.35% | 1289 | 14.58% | 12.66% | 16.51% |
| 2017 | 1479 | 7.17% | 5.85% | 8.48% | 1479 | 18.73% | 16.74% | 20.72% |

For combined high/very-high distress the 2001 rate was 13.2% (95% CI: 12.6-13.9), with lower rates but overlapping CIs for all years, except 2017-2018 where rates were greater at 14.8% (95% CI: 14.2-15.5), see Table 1. Multivariable regression confirmed that compared to 2001, the 2007, 2011 and 2014 rates were significantly lower with ORs of 0.92 (95% CI: 0.86-0.98), 0.81 (95% CI: 0.76-0.88) and 0.91 (95% CI: 0.85-0.98) respectively, whilst in 2017-18 rates were greater (ORs 1.08, 95% CI: 1.01 to 1.15), see Table 5.

**Table 5.** Two-sample comparisons of psychological distress rates in 2001 and 2017/19. Positive differences indicate an increase in prevalence in 201718 compared to 2001. Given that twelve sub-group comparisons were planned (see Table 2), to minimise the occurrences of spurious positives, the alpha value was set at 0.0042.

|  | K10 very-high |  |  |  | K10 combined high/very-high |  |  |  |
| --- | --- | --- | --- | --- | --- | --- | --- | --- |
|  | difference | p-value | 95% | CI | difference | p-value | 95% | CI |
| *Male* | 1.09 | 0.001* | 0.43 | 1.75 | 1.71 | 0.004* | 0.55 | 2.87 |
| *Female* | 0.71 | 0.067 | -0.05 | 1.47 | 0.03 | 0.963 | -1.23 | 1.29 |
| *Male* |  |  |  |  |  |  |  |  |
| 18 - 24 | 1.27 | 0.045 | 0.03 | 2.51 | 4.23 | 0.023 | 0.57 | 7.89 |
| 25 - 34 | 1.86 | 0.003* | 0.62 | 3.10 | 0.84 | 0.925 | -16.59 | 18.27 |
| 35 - 44 | 0.40 | 0.505 | -0.77 | 1.58 | 0.21 | 0.957 | -26.59 | 28.27 |
| 45 - 54 | 1.09 | 0.160 | -0.43 | 2.61 | 1.15 | 0.179 | -0.72 | 3.81 |
| 55 - 64 | 1.42 | 0.090 | -0.22 | 3.06 | 3.14 | 0.022 | 0.449 | 5.83 |
| *Female* |  |  |  |  |  |  |  |  |
| 18 - 24 | 2.37 | 0.252 | -1.70 | 6.44 | -0.96 | 0.655 | -5.06 | 3.14 |
| 25 - 34 | -1.05 | 0.149 | -2.48 | 0.379 | -1.98 | 0.133 | -4.57 | 0.61 |
| 35 - 44 | -0.16 | 0.826 | -1.59 | 1.27 | -2.32 | 0.047 | -4.61 | -0.03 |
| 45 - 54 | 0.51 | 0.995 | -183.5 | 183.4 | 0.95 | 0.462 | -1.59 | 3.49 |
| 55 - 64 | 3.63 | <0.0001* | 1.94 | 5.32 | 6.35 | <0.0001* | 3.45 | 9.33 |

In analysis by gender, very-high distress was more prevalent in women at 5.2% (95% CI: 5.0-5.4) compared to men at 3.3% (95% CI: 3.1-3.5). Combined high/very-high distress was also more prevalent in women at 16.1% (95% CI: 15.7-16.4) compared to men at 10.9% (95% CI: 10.6-13.9). Multivariable regression confirmed that women had greater odds for very-high distress (OR 1.56, 95% CI: 1.45 to 1.67) and for combined high/very-high distress (OR 1.54, 95% CI: 1.47 to 1.60), compared to men, see Tables 1 and 5.

In analysis by age groups, very-high distress rates ranged between 3.6% (95% CI: 3.3-3.8) in those aged 25-34 years to 5.4% (95% CI: 5.1-5.7) in 45-54 years. Multivariable regression showed that only the 45-54 age group had significantly greater odds for very-high distress (OR 1.56, 95% CI: 1.45 to 1.67) compared to the youngest group at 4.1% (95% CI: 3.6-4.5). Prevalence of combined high/very-high distress was greatest in those aged 18-24 years at 15.8% (95% CI: 15.0-16.6). Multivariable regression confirmed that all other age-groups had significantly lower rates than those aged 18-24 years with an OR of 0.77 (95% CI: 0.72-0.83), 0.79 (95% CI: 0.73-0.85), 0.86 (95% CI: 0.80-0.93) and 0.79 (95% CI: 0.73-0.85) for 15-34, 35-44, 45-54 and 55-64 years respectively.

Subgroup breakdowns for each survey by gender and age are in Table 2. The most marked increase in psychological distress between 2001 and 2017 is seen in women aged 55-64 years old, with very-high distress in 2001 at 3.5% (95% CI: 2.5-4.56) up to 7.2% (95% CI: 5.9-8.5%) in 2017. This doubling of prevalence was highly significant with a difference of 3.7% (z = 4.10, p<0.0001). Combined high/very-high distress also significantly increased from 12.4% (95% CI: 10.5-14.2%) in 2001 to 18.7 (95% CI: 16.7-20.7%) in 2017. This increase of prevalence was highly significant with a difference of 6.4 (z = 4.51, p<0.0001). Another almost doubling of combined high/very-high distress between 2001 and 2017 is seen in men aged 25-34 years old, with very-high distress in 2001 at 2.1% (95% CI: 1.4-2.8) up to 4.0% (95% CI: 2.9-5.1%) in 2017, which was also significant with a difference of 1.9% (z = 2.87, p=0.002).

In terms of geographical location, very-high distress was more prevalent in those residing in inner regional areas at 4.8% (95% CI: 4.4-5.1) compared to major cities at 4.1% (95% CI: 3.9-4.3). Combined high/very-high distress was also more prevalent in inner regional areas at 14.4% (95% CI: 13.8-15.0) compared to major cities at 13.1% (95% CI: 12.8-13.4). Multivariable regression confirmed that those in inner regional areas had greater odds for very-high distress (OR 1.16, 95% CI: 1.06 to 1.26) compared to capital cities; and greater odds for combined high/very-high distress (OR 1.11, 95% CI: 1.05 to 1.17) compared to capital cities, see Tables 2 and 5.

## 4. Discussion

### Key trend findings

The latest NHS reveals the highest estimates for very-high distress rates in Australia this century, at 5.1%. Relatively, there has been an increase of 34% since 2001; a modest rate of decline in the late 2000s was unsustained. Rates and changes vary between subgroups: very-high distress in women aged 55-64 has doubled this century so far, a highly significant and concerning finding. Very-high distress also increased in males, significantly in the age group 25-34. Overall, distress is greatest in women aged 18-24 years. Considering location, very-high distress is somewhat more common in inner regional Australia than elsewhere.

Given the decade 2008-2017 included major mental-health developments such as the Better Access initiative (12), we might have hoped for better population outcomes. Instead, these years – and the rest of this century to date before the COVID-19 pandemic - as measured using psychological distress in national surveys have seen the mental health of Australians get appreciably worse.

Why this happened bears investigation. A lack of data precludes analysis of service quality (10). Considering key influences on healthcare utilisation (16), candidate environmental causes which may have contributed to this include the concern that rising inequity may have driven up prevalence (5). Wealth inequity increased in Australia between 2003 and 2016, with the most affluent financial quintile experiencing a 53% increase in wealth, and the poorest, a 9% decline (14).

For women in the 55-64 age group, income stress may often be significant - among other stresses - due to structural and occupational factors such as impacts of divorce, gender pay gap, and insecure work (17). Women in this age group are more likely to be at risk of poverty and homelessness in Australia (18). Greater socioeconomic disadvantage of area (5, 19) and lower personal income (20) are associated with 2-3 fold increased prevalence of mental health issues, so these influences may be contributing to the findings here regarding increasing psychological distress in this demographic group (19). Income stress may further compromise access healthcare services which require co-payments. Services may be particularly deficient in response to key influences on mental health in this group including family violence (21).

For the first time we find significantly greater psychological distress among Australians residing in inner regional Australia compared to elsewhere. Many such areas have greater socioeconomic disadvantage and proportionally much lower mental health care use (6). In addition to understanding social determinants operating in these areas and responding to these, addressing disparities in mental healthcare outside major cities should be a priority. The recent Federal Budget provided welcome continuation for popular telehealth services, though only until December 2021

### Limitations

These data sources pre-date the COVID-19 pandemic. The impacts of COVID-19 are significant and mental health impacts have been reported to be greater in women. We note that further subgroup examination may add to our understanding of operation of other social determinants but is beyond the scope of this paper. For the NHS done to date, remote areas are out of scope. Forthcoming detailed mental health surveys will apply more specifically valid diagnostic instruments (22).

### Broad policy implications

Why have enhanced mental health services so far not achieved gains for the community when measured as a whole (5)? Why is psychological distress increasing in prevalence? Repeated inquiries have found that Australia’s mental health system is hard to navigate (23). Poor articulation of responsibilities between different levels of government have permitted the evolution of a proliferation of service structures (10, 23). There is a lack of equity in access and poor continuity of care. Comprehensive, recovery-oriented and person-centred care is very rare (5). Even recent budget announcements only go part way to bridging the gap between the level of funding and the burden of disease for which mental illness is responsible (24).

In this context, resources for mental health care are very precious and cannot be wasted. They should be carefully directed to where it they are needed most, and to whom, including with attention to equity in service provision, then to delivery of acceptable and effective kinds of help.

As we aspire to improve mental health services, and improve population mental health in an equitable way (24), we need to understand and address root causes. If inequity or other social or economic conditions are driving prevalence up, then we need models that quantify this. Perhaps such influences are so powerful that mental health services cannot reasonably be expected to influence prevalence. Effective actions must instead model broader cooperation across a mental health ‘ecosystem’, in economic, housing, educational, employment and other policy spheres across portfolios and governments. Modelling also may inform how mental health services may be adapted to achieve better outcomes for more people and influence prevalence even against such adverse conditions. We already have the expertise and tools in Australia to do the modelling that could inform such direction setting (5, 25).

This is perhaps Australia’s greatest challenge in mental health reform now, beyond the usual calls for political will and more funding. It is that we must abandon the planning strategies of the past in favour of a new and more sophisticated approach. Informed by contemporary modelling and paying particular attention to equitable implementation of evidence-based care, treatment and recovery support, we should be seeking to set and implement a broad and bold agenda, one that could provide all Australians with enjoyment of the greatest attainable standard of mental health.

## Data Availability

This study was a large-scale secondary analysis (n=78, 204) of K10 data from six national data sources collected from working aged Australian adults across the National Health Surveys (NHS) (2001-02, 2004-05, 2007-08, 2011-12, 2014-15, 2017-18), collected by the Australian Bureau of Statistics (ABS), and publicly available to access via the Microdata download portal on the ABS website: https://www.abs.gov.au/websitedbs/d3310114.nsf/home/microdatadownload

## DECLARATIONS

### Authors’ contributions

JE and GM were responsible for the conception and design of the study. JE and SD were responsible for the acquisition of data, and analysis. All authors made contributions to all of the following: (1) the interpretation of data, (2) drafting the article or revising it critically for important intellectual content, (3) approval of the version to be submitted.

## Acknowledgement

This study was made possible by the high quality data collection conducted by the Australian Bureau of Statistics.

